# Selective mutism in China: a nationwide survey and case-control study

**DOI:** 10.1101/2022.10.19.22281244

**Authors:** Xinwen Hu, Junhua Reitman, Stephen W. Pan

## Abstract

**Background:** Selective mutism (SM) is an anxiety disorder characterized by a consistent failure to speak in particular public settings despite speaking normally in other situations. However, quantitative SM research from China remains scanty at best. In response, this study aimed (1) to describe the sociodemographics and experiences of children and families with SM in China and (2) to identify sociodemographic and environmental correlates of SM in China.

**Methods:** This case-control mixed-methods study was composed of 172 cases with SM and 179 controls, recruited by online surveys. Multilevel logistic regression was performed to examine the association between potential correlates and SM.

**Results:** Only 51.2% of SM cases were diagnosed by a professional, and 31.1% of SM cases that received treatment were guided by a professional. Child SM was associated with: having two parents with an introverted personality (Adjusted odds ratio (AOR): 15.05, (95% confidence interval (CI): 5.39 – 42.07), being born to a mothers aged ≥ 35 (AOR 6.44, 95%CI: 1.24-33.43), and having a sibling (AOR 1.92, 95% CI: 1.00 – 3.70). However, child SM was inversely associated with ever receiving bilingual (AOR 0.20, 95% CI 0.10-0.39) education or being enrolled in an international school (AOR 0.12, 95% CI 0.04-0.35).

**Conclusions:** Findings suggest that many children with SM in China have not received professional treatment or interventions. Hereditary and social environmental factors may be contributing to childhood onset of SM in China. Novel policies such as access to special education resources, SM-training for therapists, and school support are needed to enhance the early detection and treatment of SM in China.

## Background

Selective mutism (SM) is an anxiety disorder characterized by a consistent failure to speak in particular public settings (e.g., school), while being able to speak normally in other situations (e.g., home) [1,2]. The disorder is listed among the anxiety disorders in the Diagnostic and Statistical Manual of Mental Disorders, 5^th^ Edition (DSM-5) [2]. To diagnose this disorder, the DSM-5 stipulates that speech inhibition should last for at least one month. The diagnosis cannot be established during the first month at school, where the absence of speech may be attributed to children’s nervousness in an unfamiliar environment instead of a mental disorder. Moreover, according to the DSM-5, the lack of speech is not the result of a lack of familiarity with or discomfort with the spoken language needed in the social situation. In addition, the failure to speak is not better explained by a communication disorder and does not occur exclusively during the course of autism spectrum disorder, schizophrenia, or another psychotic disorder [2].

The prevalence of SM among children is estimated between 0.03% and 1%, and the mean age of onset is between 2 and 5 years, thus indicating that SM is a relatively rare childhood disorder [1,2]. SM has a mean duration of 8 years, after which the total lack of speech in certain settings usually dissipates [3]. The difficulties associated with SM include speech problems, fears, and social phobia, which can interfere with children’s daily function, and educational and occupational achievement [2,4].

International literature suggests that several sociodemographic and environmental factors may be associated with SM among children. For example, Koskela et al. [5] stated that compared to a child of a mother who was married or in a relationship, the child of a single mother was twice as likely to develop SM. A telephone interview study revealed that the prevalence of SM among immigrant children was 2.2%, while the prevalence among children in native families was 0.5%, indicating that immigrant status is a potential contributing factor to SM development [6]. However, there is only limited evidence of the associations between these factors and children’s SM, and some of these findings remain controversial. For example, Elizur, Perednik [6] suggested that second language acquisition is a major correlate of the development of SM in immigrant children. However, Starke [7] argued that second language acquisition alone could not cause SM. She held the view that although bilingualism is associated with SM in immigrant children, a low orientation to the mainstream culture and anxiety are the major causes and predictors of their mute behavior [7]. The association between maternal age and offspring SM remains controversial as well. While McGrath et al. [8] suggested an association between maternal age and SM among their children, the study by Koskela et al. [5] reported no significant association between these factors.

Several other sociodemographic and environmental variables might also be associated with children’s risk for SM, including parental educational attainment and major life events such as relocation [9-11]. Nevertheless, there is a dearth of knowledge on the relationship between these factors and the onset of SM. Moreover, although Chavira et al. [12] reported that shyness was more common in parents of children with SM, it is unclear whether there is an association between introversion of parents and offspring SM.

In terms of treatment, the international literature suggests that behavioral or cognitive-behavioral interventions and pharmacotherapy with selective serotonin reuptake inhibitors are effective for treating SM [1]. Individualized treatment plans with the combined effort of teachers, parents, and clinics has been recommended to help children overcome SM [13]. However, there are meager data on how families with SM children cope with SM and how they perceive the effectiveness of SM treatments.

### Selective Mutism in China

China has the second-largest number of children in the world. In 2015, there were 271 million children under the age of 17 years in China, accounting for 20% of the China population and 15% of the world’s child population [14]. Therefore, although SM is a relatively rare condition, China likely has many children living with SM due to its large base population. Due in part to such concerns, the Selective Mutism Association of China (SMAC) (https://www.selectivemutism.org.cn/) was launched in 2018 [15]. The SMAC is a non-governmental organization composed of professional therapists and parents. The goal of the SMAC is to spread knowledge about SM, raise public awareness, and help SM patients and families receive timely SM support and treatment [15].

Children in China have potentially unique triggers for SM. Second language acquisition when receiving bilingual education or studying at an international school may be one of these triggers, as English and foreign language training for children has become more popular in recent years in China [16]. Using language dialects for communication at home is also a potential trigger. In China, there are diverse dialects and most of them are quite different from Mandarin, the official language used in public settings. Additionally, parental divorce is another potential trigger for SM, as the divorce rate in China has been rising in recent years, and parental divorce can increase children’s anxiety levels [17,18] and increase the likelihood of a child growing up in a single parent household. Moreover, it is estimated that 69 million children in China are left behind in the care of their extended family and friends as their parents move to other cities for employment [19]. Furthermore, the unique One-Child Policy implemented between 1980 and 2015 makes China the country with the highest number and percentage of only children globally [20]. Being a left-behind child and being an only child may also be a trigger for SM since parental companionship and siblings play a vital role in children’s cognitive development and mental health [21,18].

Nonetheless, there remains limited knowledge about SM in China. To the best of our knowledge, there has been virtually no quantitative SM research in China, and no systematic data on the experiences and treatment of children and families dealing with SM in China has been published.

### Rationale and objectives of the study

To better understand SM in China and its correlates, this nationwide case-control survey study aims (1) to describe the sociodemographics and experiences of children and families with SM in China and (2) to identify sociodemographic and environmental correlates of SM in China.

## Methods

### Recruitment

This case-control study was conducted in a partnership with SMAC. We recruited case participants (i.e., children suspected of ever having SM) by disseminating the online survey questionnaire in a WeChat group organized by the SMAC (WeChat is a Chinese messaging and social media application). In addition, we recruited control participants (i.e., children not suspected of ever having SM) by using the survey service offered by Wenjuanxing (WJX), a professional online questionnaire survey platform. The Wenjuanxing sample database covers more than 2.6 million respondents who register under real names and have diverse socio demographics [22,23]. The recruitment criteria for the study were (1) Currently residing in China, (2) Having at least one child currently between the ages of 2 and 17 years who currently resides in China. Before filling in the questionnaire, each participant completed an informed consent form.

Recruitment of cases started on March 28^th^, 2022 and ended on April 18^th^, 2022. Recruitment of controls began on April 13^th^, 2022 and ended on April 18^th^, 2022. Each of the study participants (i.e., parents of children) reported information for one or two children in their family. We stipulated that each family should only complete one questionnaire.

### Measures

Online questionnaires collected information about parent and child sociodemographics, and whether or not children exhibited symptoms of SM. In addition, the survey for cases included more detailed questions about each child’s SM diagnosis, severity, and impact of SM on parental and child stress.

#### SM case definition

The DSM-5 is the latest updated guideline for diagnosing mental disorders developed by the American Psychiatric Association (APA) in 2013 [2]. In our survey, we translated the DSM-5’s diagnostic criteria of SM into Chinese, and parents were asked if they felt their child met the criteria of SM. Possible responses were yes, no, and unsure.

#### Selective Mutism Questionnaire (SMQ)

The SMQ is a 17-item parent-report measure of children’s speaking behavior developed by Bergman et al. [24]. The frequency of a child’s speaking behavior is rated by the parent using a scale ranging from 0 (never), 1 (seldom), 2 (often), and 3 (always). Lower scores represent higher speech inhibition [24]. In typically developed children (i.e., children without SM) and children with SM, the mean scores were ≥ 2.5 and 0.5 [25], respectively. Based on the 17-item SMQ, Letamendi et al. [26] published a 13-item SMQ, where four other items with low interpretability or relatively high secondary cross-loadings were dropped. The 13-item SMQ had internal consistency ranging from moderate to high and good convergent and incremental validity [26]. We translated the 13-item SMQ into Chinese and used it in the survey.

In addition, the SMQ contains items regarding the stress of children’s current SM behavior for children and parents. A different scale is used for these items (0 = Not at all, 1 = Slightly, 2 = Moderately, 3 = Extremely) [24]. We used two items that assessed how stressful the child’s SM behavior was for the parent and child.

#### Parental Stress

Parental stress was measured by participants’ five-point response (from “strongly disagree” to “strongly agree”) to the statement “having children leaves little time and flexibility in my life”. This measure came from the parental stress scale, a 18-item measure of parental stress developed by Berry, Jones [27].

### Statistical analysis

To describe the sociodemographics and experiences of children and families with SM in China, a descriptive analysis was performed. In addition, a thematic analysis was done to analyze treatment experiences by coding open-ended qualitative responses and grouping the codes into themes. To identify sociodemographic and environmental correlates of SM in China, a multilevel binary logistic regression with random intercepts was performed, with individual children clustered within families. Multicollinearity was not detected among the potential correlates. The measures of association between the examined factors and SM onset were presented as bivariable odds ratios (OR) with 95% confidence intervals (CI). Two-sided *p*-values < 0.05 were considered statistically significant.

### Ethical assessment

The research protocol was approved by the University Ethics Committee of Xi’an Jiaotong-Liverpool University.

## Results

The sample included 172 cases of children suspected of ever having SM and 179 controls who were not suspected of ever having SM. Most participants were female (89.0% of cases; 79.3% of controls) and most households had two parents with a bachelor’s degree (63.4% of cases; 74.3% of controls). Parents of cases were more likely to be over 40 years old, compared to parents of controls (23.8% vs 8.4 %). Additional sociodemographic information is presented in Table 1.

**Table 1:**
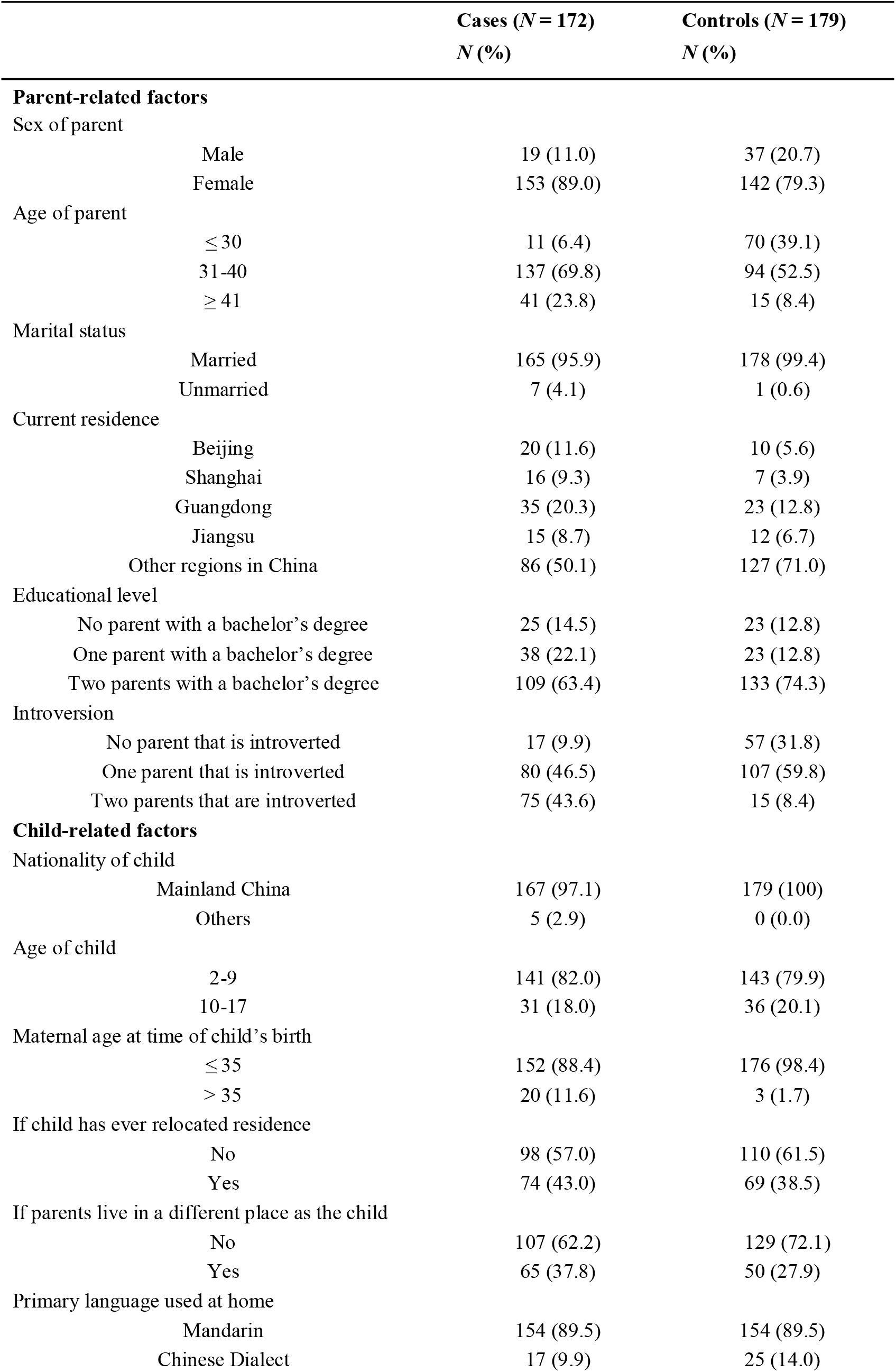
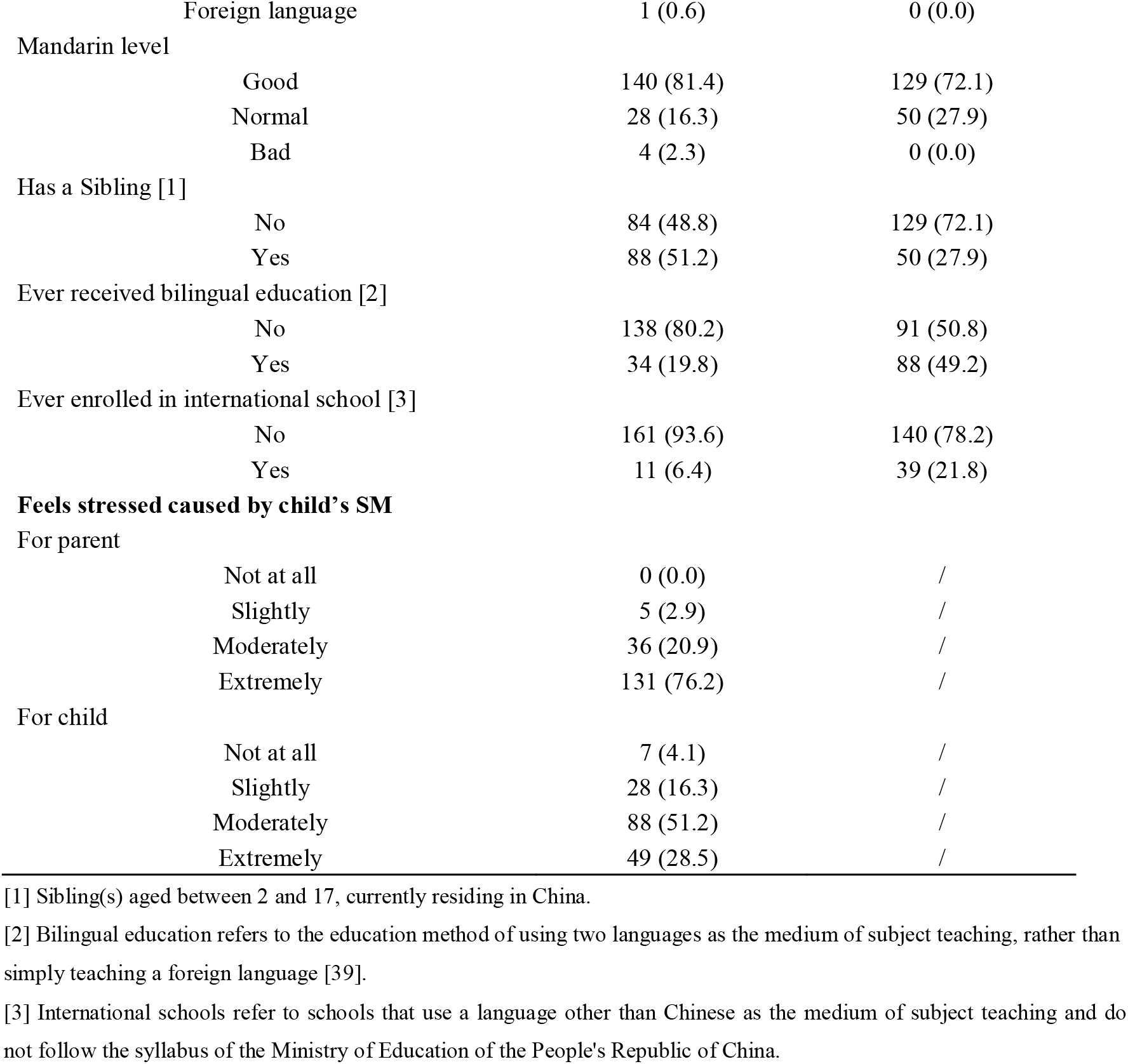
Characteristics of children and their parents

|  |  | Cases (*N* = 172)<br>*N* (%) | Controls (*N* = 179)<br>*N* (%) |
| --- | --- | --- | --- |
| **Parent-related factors** |  |  |  |
| Sex of parent |  |  |  |
|  | Male | 19 (11.0) | 37 (20.7) |
|  | Female | 153 (89.0) | 142 (79.3) |
| Age of parent |  |  |  |
|  | ≤ 30 | 11 (6.4) | 70 (39.1) |
|  | 31-40 | 137 (69.8) | 94 (52.5) |
|  | ≥ 41 | 41 (23.8) | 15 (8.4) |
| Marital status |  |  |  |
|  | Married | 165 (95.9) | 178 (99.4) |
|  | Unmarried | 7 (4.1) | 1 (0.6) |
| Current residence |  |  |  |
|  | Beijing | 20 (11.6) | 10 (5.6) |
|  | Shanghai | 16 (9.3) | 7 (3.9) |
|  | Guangdong | 35 (20.3) | 23 (12.8) |
|  | Jiangsu | 15 (8.7) | 12 (6.7) |
|  | Other regions in China | 86 (50.1) | 127 (71.0) |
| Educational level |  |  |  |
|  | No parent with a bachelor's degree | 25 (14.5) | 23 (12.8) |
|  | One parent with a bachelor's degree | 38 (22.1) | 23 (12.8) |
|  | Two parents with a bachelor's degree | 109 (63.4) | 133 (74.3) |
| Introversion |  |  |  |
|  | No parent that is introverted | 17 (9.9) | 57 (31.8) |
|  | One parent that is introverted | 80 (46.5) | 107 (59.8) |
|  | Two parents that are introverted | 75 (43.6) | 15 (8.4) |
| **Child-related factors** |  |  |  |
| Nationality of child |  |  |  |
|  | Mainland China | 167 (97.1) | 179 (100) |
|  | Others | 5 (2.9) | 0 (0.0) |
| Age of child |  |  |  |
|  | 2-9 | 141 (82.0) | 143 (79.9) |
|  | 10-17 | 31 (18.0) | 36 (20.1) |
| Maternal age at time of child's birth |  |  |  |
|  | ≤ 35 | 152 (88.4) | 176 (98.4) |
|  | > 35 | 20 (11.6) | 3 (1.7) |
| If child has ever relocated residence |  |  |  |
|  | No | 98 (57.0) | 110 (61.5) |
|  | Yes | 74 (43.0) | 69 (38.5) |
| If parents live in a different place as the child |  |  |  |
|  | No | 107 (62.2) | 129 (72.1) |
|  | Yes | 65 (37.8) | 50 (27.9) |
| Primary language used at home |  |  |  |
|  | Mandarin | 154 (89.5) | 154 (89.5) |
|  | Chinese Dialect | 17 (9.9) | 25 (14.0) |

|  |  |  |
| --- | --- | --- |
| Foreign language | 1 (0.6) | 0 (0.0) |
| Mandarin level |  |  |
| Good | 140 (81.4) | 129 (72.1) |
| Normal | 28 (16.3) | 50 (27.9) |
| Bad | 4 (2.3) | 0 (0.0) |
| Has a Sibling [1] |  |  |
| No | 84 (48.8) | 129 (72.1) |
| Yes | 88 (51.2) | 50 (27.9) |
| Ever received bilingual education [2] |  |  |
| No | 138 (80.2) | 91 (50.8) |
| Yes | 34 (19.8) | 88 (49.2) |
| Ever enrolled in international school [3] |  |  |
| No | 161 (93.6) | 140 (78.2) |
| Yes | 11 (6.4) | 39 (21.8) |
| **Feels stressed caused by child's SM** |  |  |
| For parent |  |  |
| Not at all | 0 (0.0) | / |
| Slightly | 5 (2.9) | / |
| Moderately | 36 (20.9) | / |
| Extremely | 131 (76.2) | / |
| For child |  |  |
| Not at all | 7 (4.1) | / |
| Slightly | 28 (16.3) | / |
| Moderately | 88 (51.2) | / |
| Extremely | 49 (28.5) | / |

[1] Sibling(s) aged between 2 and 17, currently residing in China.
[2] Bilingual education refers to the education method of using two languages as the medium of subject teaching, rather than simply teaching a foreign language [39].
[3] International schools refer to schools that use a language other than Chinese as the medium of subject teaching and do not follow the syllabus of the Ministry of Education of the People's Republic of China.

### SM severity, treatment, and impact

Among 172 case children with suspected SM, 51.2% were diagnosed by a professional and 167 (97.1%) had symptoms that had been detected by the participants. The mean values, standard deviations, and ranges of the mean scores of the 13-item SMQ were 1.18 ± 0.41 (0.15-2.46) among all 172 case children and were 1.16 ± 0.40 (0.15-2.00) among 78 case children who had not received treatment.

Among 167 children whose symptoms had been previously detected by participants, 89 (53.3%) had received treatment; 52 (31.1%) had received treatment under the instruction of a professional; the percentages of children who received very effective, effective, and ineffective professional treatment based on parents’ self-assessment were 20.2%, 53.9%, and 25.8%. Based on the thematic analysis of answers to an open-ended question regarding SM children and their families’ treatment experiences (see Table 2 for the illustrative quotes), it appears that few educational institutions in China provide special support for children with SM. In addition, some participants indicated that there is a lack of professionals experienced with SM. Among participants whose child had received professional SM treatment, multiple participants cited the use of Cognitive-Behavioral Therapy, which they regarded as an effective method. Sandplay therapy, which is a psychotherapy method where clients are asked to play in a specially proportioned sandbox [28], was another popular treatment method for SM among the participants. Sandplay therapy was reported to be expensive and have little to no effect on their child’s SM.

**Table 2:**
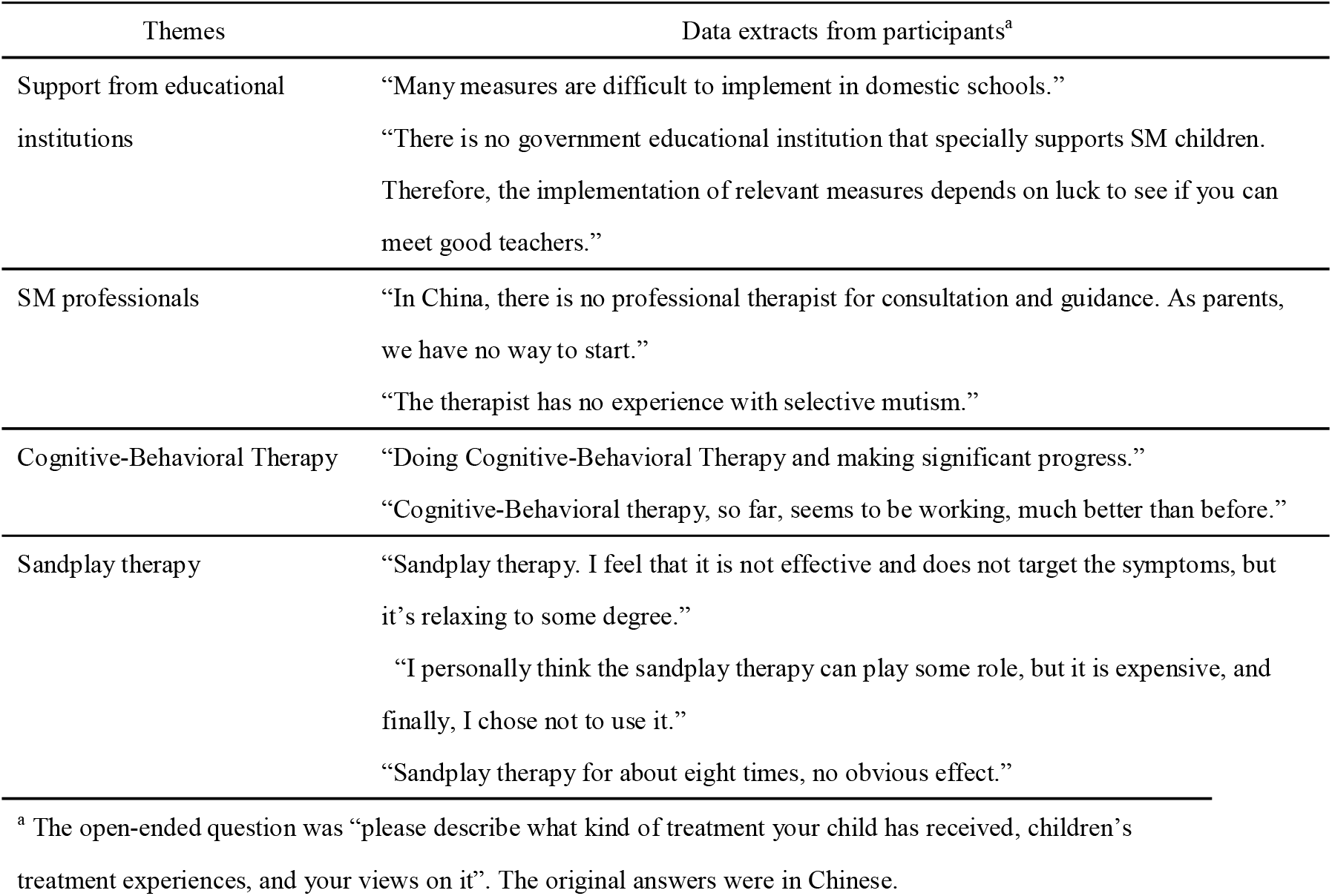
Thematic analysis on treatment experiences

| Themes | Data extracts from participants<sup>a</sup> |
| --- | --- |
| Support from educational institutions | “Many measures are difficult to implement in domestic schools.”<br>“There is no government educational institution that specially supports SM children. Therefore, the implementation of relevant measures depends on luck to see if you can meet good teachers.” |
| SM professionals | “In China, there is no professional therapist for consultation and guidance. As parents, we have no way to start.”<br>“The therapist has no experience with selective mutism.” |
| Cognitive-Behavioral Therapy | “Doing Cognitive-Behavioral Therapy and making significant progress.”<br>“Cognitive-Behavioral therapy, so far, seems to be working, much better than before.” |
| Sandplay therapy | “Sandplay therapy. I feel that it is not effective and does not target the symptoms, but it’s relaxing to some degree.”<br>“I personally think the sandplay therapy can play some role, but it is expensive, and finally, I chose not to use it.”<br>“Sandplay therapy for about eight times, no obvious effect.” |

<sup>a</sup> The open-ended question was “please describe what kind of treatment your child has received, children’s treatment experiences, and your views on it”. The original answers were in Chinese.

In terms of SM as a source of stress, the parents of 167 SM children (97.1%) reported themselves as being moderately or extremely stressed by their child’s SM; while 137 (79.7%) reported that their children were moderately or extremely stressed by SM (Table 1). Compared to parents of children without SM, the parents of SM children were more likely to agree or strongly agree that having children leaves little time and flexibility in their lives (50.6% vs 35.8%).

### Correlates of SM

Table 3 presents unadjusted and adjusted measures of association between SM and select correlates. Results of adjusted multilevel regression analysis indicated that compared to children with no introverted parent, children of two parents with an introverted personality had greater odds of having SM (Adjusted Odds Ratio (AOR) 15.05, 95% Confidence Interval (CI) 5.39-42.07). Children born to mothers aged > 35 had significantly greater odds of SM (AOR 6.44, 95%CI 1.24-33.43). Children with a sibling had significantly greater odds of SM (AOR 1.92, 95%CI, 1.00-3.70) than those without a sibling. Children who had ever received bilingual (AOR 0.20, 95% CI 0.10-0.39) education or enrolled in an international school (AOR 0.12, 95% CI 0.04-0.35) had lower odds of SM than those who were not.

**Table 3:** Participant characteristics and correlates of childhood selective mutism

|  | Unadjusted odds ratios<br>(95% Confidence intervals) | Adjusted odds ratios<br>(95% Confidence intervals) |
| --- | --- | --- |
| **Parent-related factors** |  |  |
| Marital status |  |  |
| Married | 1.0 | 1.0 |
| Unmarried | 7.68 (0.85-69.0) | 17.46 (1.27-239.46) * |
| Educational level |  |  |
| No parent with a bachelor's degree | 1.0 | 1.0 |
| One parent with a bachelor's degree | 1.45 (0.61-3.42) | 1.78 (0.57-5.58) |
| Two parents with a bachelor's degree | 0.71 (0.36-1.43) | 1.28 (0.50-3.28) |
| Introversion |  |  |
| No parent that is introverted | 1.0 | 1.0 |
| One parent that is introverted | 2.55 (1.30-5.02) ** | 1.96 (0.87-4.40) |
| Two parents that are introverted | 18.17 (7.80-42.31) *** | 15.05 (5.39-42.07) *** |
| **Child-related factors** |  |  |
| Nationality of child |  |  |
| Mainland China | 1.0 | 1.0 |
| Others | / | / |
| Maternal age at time of child's birth |  |  |
| ≤ 35 | 1.0 | 1.0 |
| > 35 | 7.34 (2.02-26.64) ** | 6.44 (1.24-33.43) * |
| If child has ever relocated residence |  |  |
| No | 1.0 | 1.0 |
| Yes | 0.83 (0.52-1.33) | 1.28 (0.68-2.41) |
| If parents live in a different place as the child |  |  |
| No | 1.0 | 1.0 |
| Yes | 1.63 (0.99-2.69) | 1.45 (0.74-2.86) |
| Primary language used at home |  |  |
| Mandarin | 1.0 | 1.0 |
| Chinese Dialect | 0.69 (0.34-1.42) | 0.64 (0.25-1.60) |
| Foreign language | / | / |
| Has a Sibling [1] |  |  |
| No | 1.0 | 1.0 |
| Yes | 2.98 (1.80-4.95) *** | 1.92 (1.0-3.70) * |
| Ever received bilingual education [2] |  |  |
| No | 1.0 | 1.0 |
| Yes | 0.25 (0.15-0.42) *** | 0.20 (0.10-0.39) *** |
| Ever enrolled in international school [3] |  |  |
| No | 1.0 | 1.0 |
| Yes | 0.24 (0.11-0.52) *** | 0.12 (0.04-0.35) *** |

$p < 0.05^*$ , $p < 0.01^{**}$ , $p < 0.001^{***}$
[1] Sibling(s) aged between 2 and 17, currently residing in China.
[2] Bilingual education refers to the education method of using two languages as the medium of subject teaching, rather than simply teaching a foreign language.
[3] International schools refer to schools that use a language other than Chinese as the medium of subject teaching and do not follow the syllabus of the Ministry of Education of the People's Republic of China.

## Discussion

To the best of our knowledge, this is the first quantitative study to describe the sociodemographics and experiences of children and families with SM in China. Our findings revealed that only about half of SM cases were diagnosed by a professional, and less than one in three SM cases received treatment guided by a professional. This could be partly due to a paucity of both trained professionals experienced with SM and special support from educational institutions. Although children with special needs have been increasingly educated in mainstream classrooms within China, mainstream public schools often lack the resources and organization needed to meet such students’ needs [29]. The relatively large size of classrooms (often ≥40 students in urban settings) [30] constrains the time regular classroom teachers can devote to students with special needs. Teachers tasked with supporting students with special needs often have limited special education training and may need to take on such duties in addition to other job duties, but without additional financial compensation [31]. In more developed regions, private professional therapists can be hired to implement SM treatment programs in school settings, but may be prohibitively expensive for many if not most families. Due to the fact that many children with SM do not exhibit any intellectual disability, there is a very real risk that educators in mainstream schools may incorrectly attribute SM behavior to shyness or introversion, rather than a recognized anxiety disorder [32]. In fact, anecdotal evidence from the SMAC suggests that many children in China living with SM demonstrate relatively high academic achievement. Greater access to special education resources and SM interventions are needed in China.

Results from our study also indicated that SM is a substantial source of stress for both parents and their children in China. Part of this stress may be exacerbated by parent-teacher relations, wherein parents are expected to be personally engaged with their child’s studies and provide at-home support as the teacher requests [30]. It is conceivable that homework and at-home “make-up” assignments may be greater for students with SM that are unable to participate in school activities that require verbal communication. Given that speech becomes increasingly salient for developing social bonds and communication skills as the child ages, it is likely that SM-attributed stress will only increase over time unless SM symptoms are resolved. In the interest of the well-being of the child and their parents, it is imperative that SM treatment begin as soon as possible for the child [32-34]. Chinese language resources for therapists and parents of children with SM are growing, but have been relatively sparse to date with a few exceptions [35].

Previous research from outside Asia has suggested that genetics and parental personality has a strong influence on SM onset [1]. Findings from our sample from China corroborate these previous studies, with SM cases having significantly greater odds of two introverted parents. However, contrary to our original expectations, having a sibling was positively associated with SM. This association could be confounded by unmeasured effects of socioeconomic status, but could not be assessed in the present study. As more families have multiple children following the end of the “one child policy,” future research should examine how sibling dynamics may or may not be influencing SM onset in China.

Advanced maternal age was strongly associated with SM in the current study, but we can only speculate upon the mechanism of association. A large nationwide study from Finland found advanced maternal age to be significantly associated with SM in unadjusted models, but was no longer significantly associated with SM after psychopathology of parents was controlled for Koskela et al. [5]. It is conceivable that parental psychopathologies can be confounding the association between advanced maternal age and child SM, but it is also possible that maternal psychopathologies are mediating the effects of advanced maternal age on child SM [36]. Longitudinal studies are needed to disentangle the relationship between maternal age, maternal psychopathologies, and SM in offspring.

In contrast to the earlier research by Elizur, Perednik [6], we found an inverse association between bilingual education and SM. Nearly all of our samples (98.6%) were of Chinese nationality and resided in China. Therefore, the cross-cultural conflict they experienced in daily life might be limited. As suggested by previous research, cross-cultural conflict experienced by immigrant children could be one of the main causes of their mute behavior and the mediator of the association between bilingualism and SM among them [7]. Similarly, we also found that children who ever enrolled in an international school in China had lower odds of SM. However, the association between bilingual education and international school and SM might be underestimated given that the confounding effect of family income was not controlled. According to Koskela et al. [5], people of lowest social economic status were more likely to have children with SM. Furthermore, children from low-income family are generally less likely to go to bilingual and international school due to the high costs.

### Limitations

The study has several limitations. Firstly, although case-control studies are efficient for studying rare conditions, there are several drawbacks inherent to this design. Firstly, because of its retrospective nature, case-control studies can only be used to identify associations between exposures and outcomes instead of the causation between them [37,38]. Therefore, future studies (ideally longitudinal cohort studies) are required to investigate the causal relationships between the identified correlates and children’s SM. The potential for recall bias is another serious weakness of case-control designs, and we are unable to rule out potential differential exposure misclassification. It is possible that parents of SM cases were more likely to over report known exposures of SM.

Selection bias also could not be ruled out. For example, a majority of participants were female and had a Bachelor’s or above degree, and over one in three participants resided in Beijing, Shanghai, Guangzhou, and Jiangsu, the most economically developed regions in China. Hence, in our samples, those who were male and had a relatively lower socioeconomic status might be under-represented. This may have caused an overestimate of SM children receiving treatment, as children from higher socioeconomic status families usually have relatively greater access to fee-based medical and social services. In addition, self-selection bias should also be noted. Parents of children with serious SM might opt out or drop out of the survey due to greater time constraints. This could result in children with more severe SM and their families being underrepresented. Future SM studies in China should attempt to recruit samples across more diverse sociodemographic subgroups.

## Conclusions

Access to SM treatment in China remains low, and is likely driven by lack of awareness and insufficient resources at medical and educational institutions. This is problematic because SM has adverse implications for childhood development and appears to be a concerning source of stress for the child and their parents. Given the severe dearth of SM research from China, it is incumbent for Chinese researchers, educators, clinicians, and policy makers to collaboratively develop novel approaches to more effectively detect, and treat SM.

## Data Availability

Data are not available.

## Acknowledgements

Many thanks to the participants for making this study possible. Much appreciation also to Yvonne Lu for her feedback on an earlier version of this manuscript. Funding for this study was provided by Xi’an Jiaotong-Liverpool University.

